# Association of inferior division MCA stroke location with populations with atrial fibrillation incidence

**DOI:** 10.1101/2021.12.06.21267371

**Authors:** Ganghyun Kim, Emilia Vitti, Melissa D. Stockbridge, Argye E. Hillis, Andreia V. Faria

## Abstract

**Background and Aim:** Anatomical features of Middle Cerebral Artery (MCA) bifurcation favors larger emboli entering the inferior rather than superior division. As cardiac source emboli are on average larger than arterial source emboli, we hypothesize that patients with atrial fibrillation have infarcts that involve more temporal and parietal lobes than frontal lobes.

**Methods:** From 1,849 patients admitted with evidence of acute or early subacute ischemic stroke on diffusion-weighted MRI (DWI), 482 affected exclusively the “lobar” MCA (sparing the lenticulostriates). They were classified as “frontal predominant” (n=105) or “temporoparietal predominant” (n=197) if at least 75% of the infarct affected the frontal lobe or the temporal and parietal lobes, respectively. Differences between stroke location (frontal or temporoparietal), sex, and race were analyzed by Chi-square test. Results: Men were more likely than women, and white people were more likely than black people to have temporoparietal strokes versus frontal strokes. Patients with confirmed diagnosis of atrial fibrillation have more temporoparietal strokes, compared to frontal strokes.

**Conclusion:** MCA ischemic strokes occur more often in temporoparietal areas in men and in white patients, populations with known elevated incidence of atrial fibrillation. Patients with confirmed diagnosis of atrial fibrillation have more temporoparietal strokes, compared to frontal strokes. Results align with the hypothesis that large emboli (mostly from cardiac source) are more likely to cause temporoparietal strokes in the MCA territory, compared to frontal strokes. This association can help guide search for the most likely etiology of infarcts.

## 1 Introduction

Previous studies have reported that Wernicke’s aphasia (due to left temporoparietal stroke) is more often due to cardioembolic stroke^1, 2^. This association was explained by a gravitational model, based on 3D volume rendering CT angiography (CTA) in 103 patients that showed inferior division lumen diameter was larger than superior division. The angle between the MCA trunk and superior division was less severe than the angle between the MCA trunk and inferior division^3^, and the takeoff of the superior division was more downward relative gravity, compared to the takeoff of the superior division. Thus, anatomical features of the MCA bifurcation favor larger emboli entering the inferior division, with larger vessel diameter, more linear path, and greater gravitational predilection, rather than superior division^3^. Based on these data, we hypothesized that strokes due to atrial fibrillation more often involve the inferior division MCA territory (temporal and parietal lobes) than superior division MCA territory (frontal lobe) in both hemispheres, in a large set of acute ischemic strokes with well-characterized lesions on MRI (n=1849). Confirmation of this hypothesis would help prioritize search for cardioembolic source in patients with inferior vision MCA ischemic stroke and provide the basis for educating patients with atrial fibrillation about non-motor signs of stroke.

## 2 Methods

This study included MRIs of patients admitted to the Comprehensive Stroke Center at Johns Hopkins Hospital with the clinical diagnosis of ischemic stroke, between 2009 and 2019 (Flowchart for data inclusion in Figure 1). It utilizes data from an anonymized dataset, created under waiver of informed consent (IRB00228775). We have complied with all relevant ethical regulations and the guidelines of the Johns Hopkins Institutional Review Board, that approved the present study (IRB00290649).

**Figure 1.**
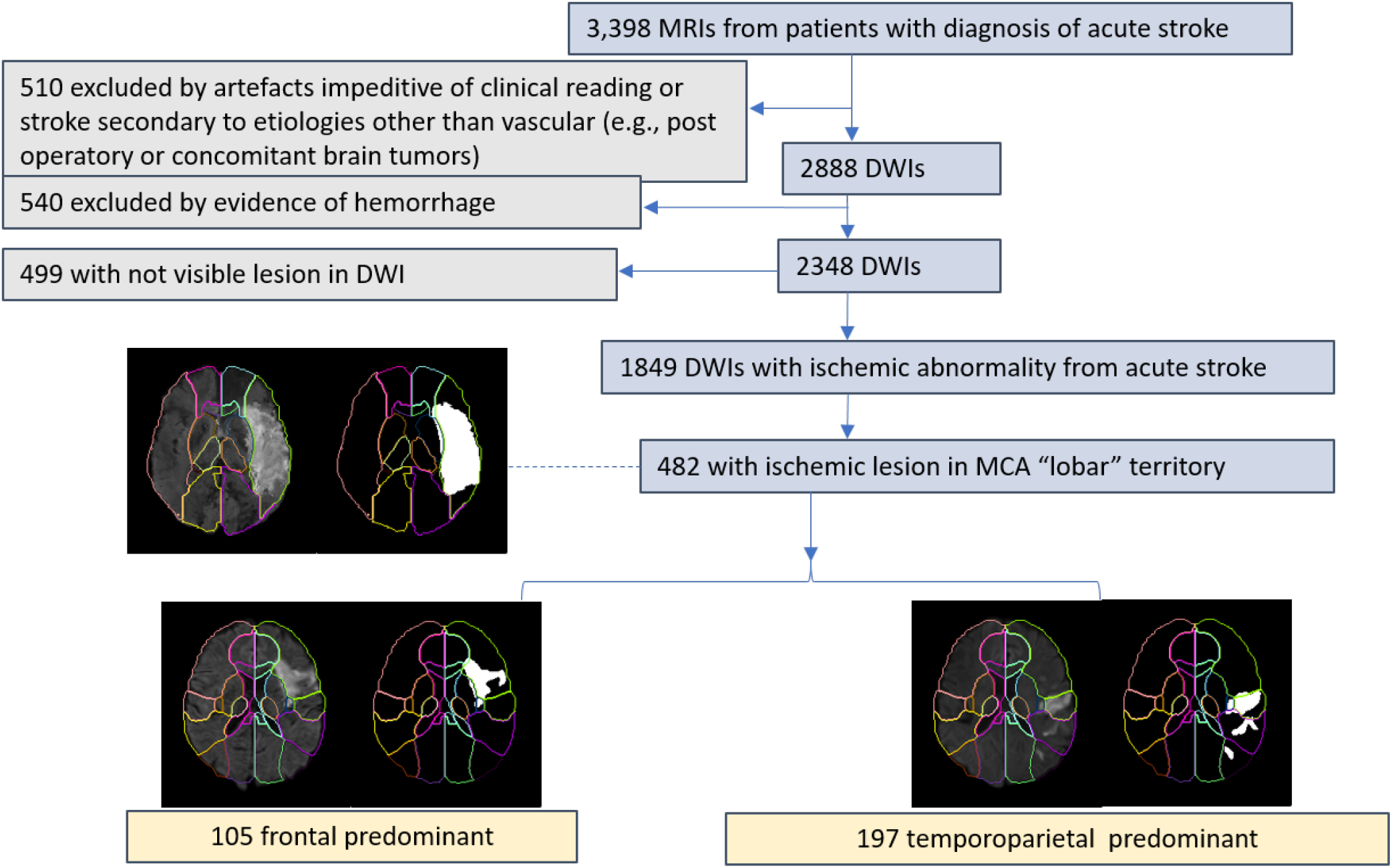
Flowchart of data inclusion

From the 2,888 DWIs quality-controlled for clinical analysis, 1,849 DWIs showed lesions classified by a neuroradiologist as result of acute or early subacute ischemic stroke, with no evidence of hemorrhage. From those, we included 482 classified as “frontal predominant” (n=105) if at least 75% of the infarct lesion was in the frontal lobe, or “temporoparietal predominant” (n=197), if at least 75% of the infarct lesion was in the temporal or parietal lobes. The lesion core was defined in DWI, in combination with the Apparent Diffusion Coefficient maps (ADC) by two experienced evaluators and was revised by a neuroradiologist until reaching a final decision by consensus. Further details are in our previous publication^4^.

The summary of demographics and lesion profiles is in Table 1. Differences between infarct location (frontal or temporoparietal), sex, and race were analyzed by Chi-square test. Race is defined here according to the Revisions to the Standards for the Classification of Federal Data on Race and Ethnicity^5^. By this classification, “white” is a person having origins in any of the original peoples of Europe, the Middle East, or North Africa and “black or African American” is a person having origins in any of the Black racial groups of Africa. In a secondary, exploratory analysis, we reviewed the patient’s records to find whether the clinical diagnosis of atrial fibrillation was confirmed.

**Table 1.** Demographic and lesion characteristics. Continuous variables are shown as mean (standard deviation).

|  | Total (N=302) | Frontal (N=105) | Temporoparietal (N=197) |
| --- | --- | --- | --- |
| Age in years | 63.3 (14.8) | 64.4 (14.5) | 62.7 (14.9) |
| Sex (female / male) | 146 (48.3%) / 156 (51.7%) | 64 (61.0%) / 41 (39.0%) | 82 (41.6%) / 115 (58.4%) |
| Race (black / white) | 155 (51.3%) / 147 (48.7%) | 62 (59.0%) / 43 (41.0%) | 93 (47.2%) / 104 (52.8%) |
| lesion volume in ml | 23.3 (39.6) | 11.9 (16.6) | 29.3 (46.4) |

## 3 Results

As shown in Table 1, there was no difference in patients’ age between frontal and temporoparietal strokes. Temporoparietal infarcts were significantly larger than frontal. The group with frontal infarcts was composed predominantly by women and black patients. The statistical comparison (Figure 2) revealed that male patients were more likely than female patients to have temporoparietal strokes versus frontal strokes and that white patients were more likely than black patients to have temporoparietal strokes versus frontal strokes (p-value 0.00613). From the 85 patients with confirmed diagnosis of atrial fibrillation, the majority (55; 65%) had temporoparietal strokes, compared to 30 (35%) with frontal strokes.

**Figure 2.**
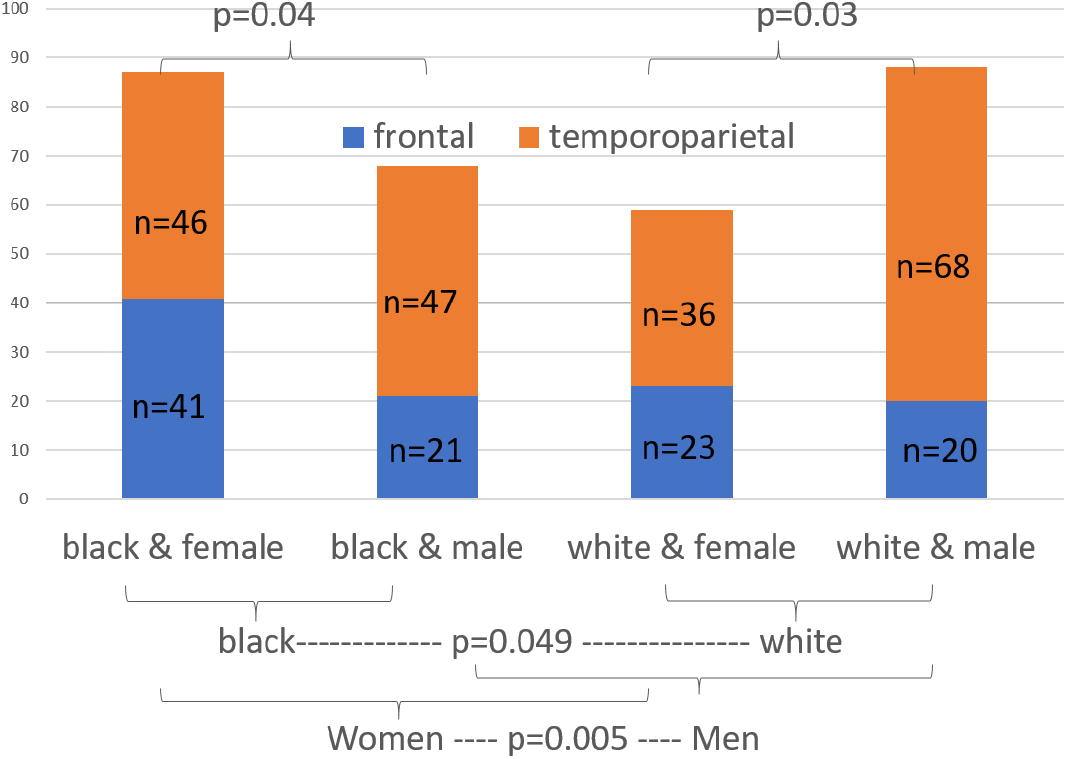
Incidence of MCA strokes in frontal and temporoparietal areas, according to patient sex and race. P shows the p-values of the Chi-square test between groups

## 4 Discussion

In a relatively large set of patients with acute ischemic stroke with well characterized lesions on MRI, we found that populations with higher incidence of atrial fibrillation (elderly male and white people^6, 7^) are more likely to have temporoparietal MCA strokes, rather than frontal MCA. We also confirmed a previous association observed in left hemisphere stroke patients^3^, which has important clinical implications. For example, posterior (temporoparietal) infarcts should raise the suspicion for a cardioembolic source of stroke, and might indicate need for longer monitoring for atrial fibrillation or transesophageal echocardiogram in patients with appropriate risk factors.

Secondly, it is important to note that the symptoms of temporoparietal infarcts are often less obvious (especially to the patient) than symptoms of frontal infarcts. While Wernicke’s aphasia may be very noticeable to listeners, the patient himself/herself might not be aware of the deficits in comprehension and meaningful speech. Moreover, left temporoparietal strokes may result in alexia, agraphia, acalculia, or right-left confusion. Thus, patients with atrial fibrillation should be educated that they are at risk for strokes without motor symptoms. Even more of a concern is that right temporoparietal strokes may result in deficits that are difficult to detect without careful assessment, such as impaired empathy^8^, impaired recognition of emotional tone of voice or facial expressions, and altered discourse and appreciation of humor and metaphor^9–13^. Even hemispatial neglect is often not detected in patients with right hemisphere stroke^14^. These patients often have anosognosia for even obvious deficits^15^.

While we did not obtain independent evidence for the gravitational mechanism underlying this association proposed by Liebeskind and colleagues, the anatomical differences the observed in bifurcation provide a plausible account. That is the larger vessel diameter, more linear path, and greater gravitational predilection in the inferior versus superior division MCA may explain why large cardiac emboli are more likely to cause temporoparietal than frontal strokes.

## Data Availability

The data that supports this study is available in Zenodo https://doi.org/10.5281/zenodo.5722425

## Data Availability

The data that supports this study is available in Zenodo https://doi.org/10.5281/zenodo.5722425^16^

## Author contribution

GK and AVF collected and analyzed the data. EV collected part of the data and drafted the work. MS significantly reviewed the draft. AEH and AVF conceived and designed the study, interpreted the data, and drafted the work.

